# Evaluating Diabetes-Specific Meal Replacements for Glycaemic Control in Overweight and Obese T2DM Patients: A Protocol for a Randomized Controlled Trial

**DOI:** 10.1101/2025.01.20.25320837

**Authors:** Wan Ain Nadirah Che Wan Mansor, Suzana Shahar, Laili Md Tohit, Roslee Rajikan, Munirah Ismail

**Author notes:** **Correspondence author:** Dr Munirah Ismail.

## Abstract

**Introduction:** Uncontrolled Type 2 diabetes mellitus (T2DM) causes micro- and macrovascular issues that hike healthcare costs and threaten global health. Previous studies suggested meal replacement (MR) therapy for T2DM, but there were inconsistencies in the results. Thus, an RCT is proposed to determine the efficacy of a diabetes-specific MR product on weight loss, glycaemic control, satiety, quality of life, metabolic gene expression, and cost-benefit in overweight and obese T2DM patients.

**Methods and analysis:** 164 overweight and obese T2DM participants will be randomly assigned to either an intervention group (n = 82) or a control group (n = 82). All participants will receive dietary counselling, however only the intervention group will receive MR for 12 weeks. Glycaemic control and obesity indicators are the study’s main outcomes. Secondary outcomes include cardiovascular disease risk marker, metabolic risk, metabolic gene profile analysis, dietary data, physical activity, satiety level, quality of life, and cost analysis. Baseline data will include socio-demographics, anthropometry, blood pressure, diet, physical activity, satiety level, quality of life, blood profile, metabolic gene expression profile, and cost-benefit analysis. Follow-up is planned at intervention weeks 6 and 12. Week 6 will assess only anthropometry, blood pressure, diet, physical activity, and satiety level. For compliance assessment, intervention group participants will bring their MR container. Week 12 will measure the same baseline parameter except socio-demographic data. Individuals that consume less than 80% of the MR will be deemed non-compliant. All parameter modifications will be documented and analysed for comparison. All statistical analyses will be conducted using IBM SPSS version 29.0 software, with a significance level of p<0.05. The cost-benefit analysis will be determined using the net present value.

**Ethics and dissemination:** This research protocol was approved by the Ethical Committee National University of Malaysia (JEP-2024-695) and registered on International Standard Randomised Controlled Trial Number (ISRCTN57040303).

**STRENGTHS AND LIMITATION OF THIS STUDY:**

- This study addresses multiple outcomes related to T2DM management, including weight reduction, glycaemic control, blood pressure, satiety level, quality of life, and metabolic gene expression, providing a holistic view of the intervention’s efficacy.
- Incorporating a cost-benefit analysis will add on a practical value by evaluating the economic feasibility of the intervention, which is essential for real-world application and healthcare policy-making.
- Since this study will investigate the effect of the partial diabetes-specific meal replacement on changes in metabolic gene expression, it will provide insights into the molecular mechanisms underlying the intervention’s effects.
- A primary limitation of the study is the absence of a placebo group and the fact it is not double blinded
- This trial is a single-centred study that is limited to the Malaysian T2DM population that visit the public health sector.

## INTRODUCTION

Obesity and type 2 diabetes mellitus (T2DM) are steadily rising in prevalence globally. Obesity exacerbates the influence of genetic vulnerability and environmental factors on the development of T2DM, as they share significant genetic and environmental characteristics in their pathogenesis. The increasing prevalence of T2DM is associated with micro- and macrovascular complications that results in increased healthcare costs, making it a public health burden worldwide, including Malaysia [1,2]. In Malaysia, 15.6% of adults have been diagnosed with diabetes, with only 44% of them have their blood sugar under control [3]. Among the various challenges faced by patients with T2DM, the primary challenge remains achieving and maintaining optimal glycaemic control [4].

Overweight or obese individuals have a higher risk of developing T2DM, and the frequent co-occurrence of both is termed ‘diabesity’ [5]. This condition is believed to be driven by chronic inflammation in obesity and highlights the shared insulin action deficiencies in both disorders [6]. A systematic review indicated that more than 30% of T2DM patients were obese across 38 of 44 trials, whereas a study conducted in Malaysia revealed that 91% of diabetic patients were classified as obese or overweight [7,8]

A balanced and well-structured diet plays a critical role in managing T2DM in order to achieve glycaemic and weight control [9]. The Diabetes Prevention Program Research Group indicated that, over the long term, an intensive lifestyle intervention (low-calorie diet and modest physical exercise) resulting in weight reduction might reduce the incidence of T2DM in overweight or obese individuals and improve impaired glucose tolerance by 27–58% [10]. Weight loss can be challenging due to the weight-gaining effects of anti-diabetic medications, as well as barriers to long-term lifestyle changes, including inadequate dietary education, reduced motivation, delays in medical follow-up, and limited family support [11]. Though implementing dietary intervention proves effective in attaining desired weight and glycaemic control among T2DM patients, this strategy can pose a great challenge for both patients and healthcare providers as it relies on patient compliance with the suggested lifestyle modification [12]

Meal replacement (MR) therapy has been recognized as one of the strategies to control weight and blood sugar levels among T2DM patients [13]. Additionally, MR has also been suggested as a strategy to help in lowering the cardiometabolic risk factor and reduce the risk for complications in T2DM [14]. MR refers to pre-packaged or commercially available food products or drinks used to replace one or more meals [15]. Most of the diabetes-specific MR are lower in glycaemic index (GI), high in fibre and lower in sugar. These foods slow gastric emptying, resulting in a slower rate of glucose absorption and a gradual release of glucose into the bloodstream. This helps regulate blood sugar levels, reduce insulin spikes, reduce food intake, and promote weight loss [16].

Studies have found that partial or complete MR can be an effective extended strategy of medical nutrition therapy in T2DM management [17,18]. However, the study’s outcome is influenced by the duration of the intervention [18–20], type of intervention [21,22], type of MR [23]and frequency of the MR given [24], which play a role in the outcome of the study. Additionally, a study among pre-diabetes and overweight patients found that low GI MR increased hormone satiety, and lowered the hunger hormone after 2 months of intervention [25].

MR can mitigate the risk of problems linked to T2DM, including diabetic foot ulcers and retinopathy, by assisting in the regulation of blood sugar levels, facilitating weight loss, ensuring adequate nutrition, and enhancing dietary compliance. This may subsequently enhance the quality of life by lowering the functional limits imposed by these issues. Variations in study protocols and routine dietary advice exist because T2DM management guidelines differ by country based on local staple foods. The latest study in Malaysia had been focusing on the effect of the MR that contain cinnamon extract among obese and overweight T2DM patients towards the hormone level [13]. However, there hasn’t been extensive exploration of the impact of MR that contain chromium on metabolic gene expression. Therefore, this study will determine the efficacy and cost benefit analysis of partial diabetes-specific MR therapy on weight reduction, glycaemic control, physical activity, quality of life and metabolic gene expression among overweight and obese T2DM patients.

## METHODS

### Study Design and Settings

This study is a two armed, randomised controlled trial. The data collection will occur at Primer and Warga Clinic, Hospital Canselor Tuanku Muhriz (HCTM) of the National University of Malaysia. Participants will be recruited beginning on December 1, 2024, with the aim of completing recruitment by April 1, 2025. The intervention and final evaluation will end on July 31, 2025. The flow of the study is mentioned in Figure 1.

**Figure 1.**
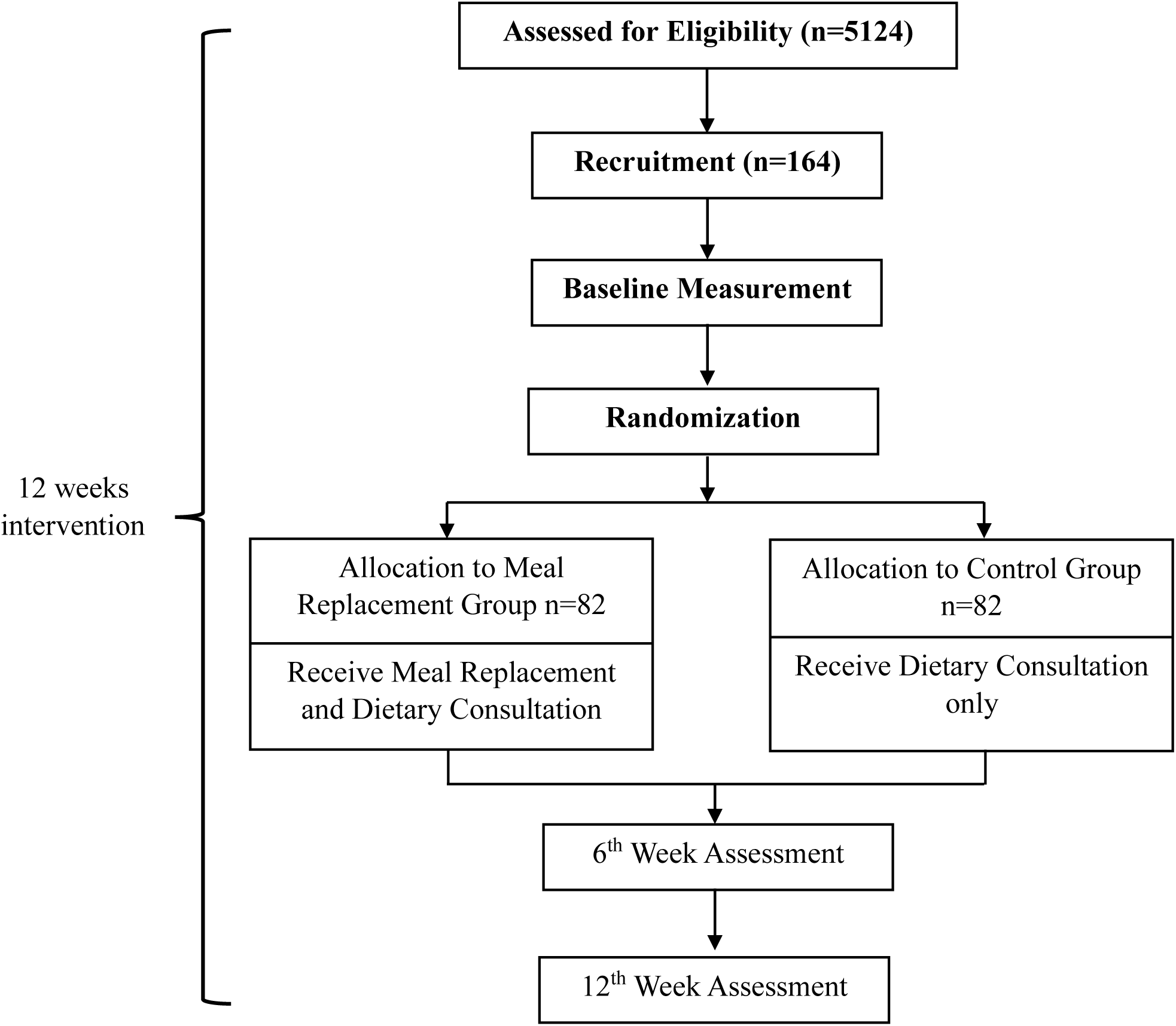
**Experimental Protocol.**

### Eligibility criteria

164 Malaysian aged between 30 and 59 years old that have been diagnosed with T2DM for at least 6 months, with baseline haemoglobin A1C (HbA1c) levels above 7.5% to 12% [13], a BMI above 23 kg/m^2^ [26] and able to communicate and understand in English or Malay language will be included in the study.

Patients are excluded from the study if he/she is diagnosed with chronic kidney diseases (GFR < 30 MI/min/1.73), hepatic diseases (ALT > 120 IU/L), cancer; pregnancy and lactating women; consuming any weight reduction products, slimming prescriptions, meal replacement; on hormone replacement therapy treatment; on treatment for cancer; involved in any clinical trial and weight loss program.

### Participants Recruitment and Randomisation

Based on inclusion and exclusion criteria, purposive sampling will determine participants. Electronic medical records databases will determine eligibility. Potential participants will be recruited at medical appointments. Details on the study will be given. For eligible participants, consent will be taken, and they will be randomly allocated to the intervention or control group. Using the minimisation approach in R, a third-party statistician will randomise by age, gender, and HbA1c during screening. Table 1 below shows a summary of parameter that will be taken during the study based on primary and secondary outcome.

**Table 1.**
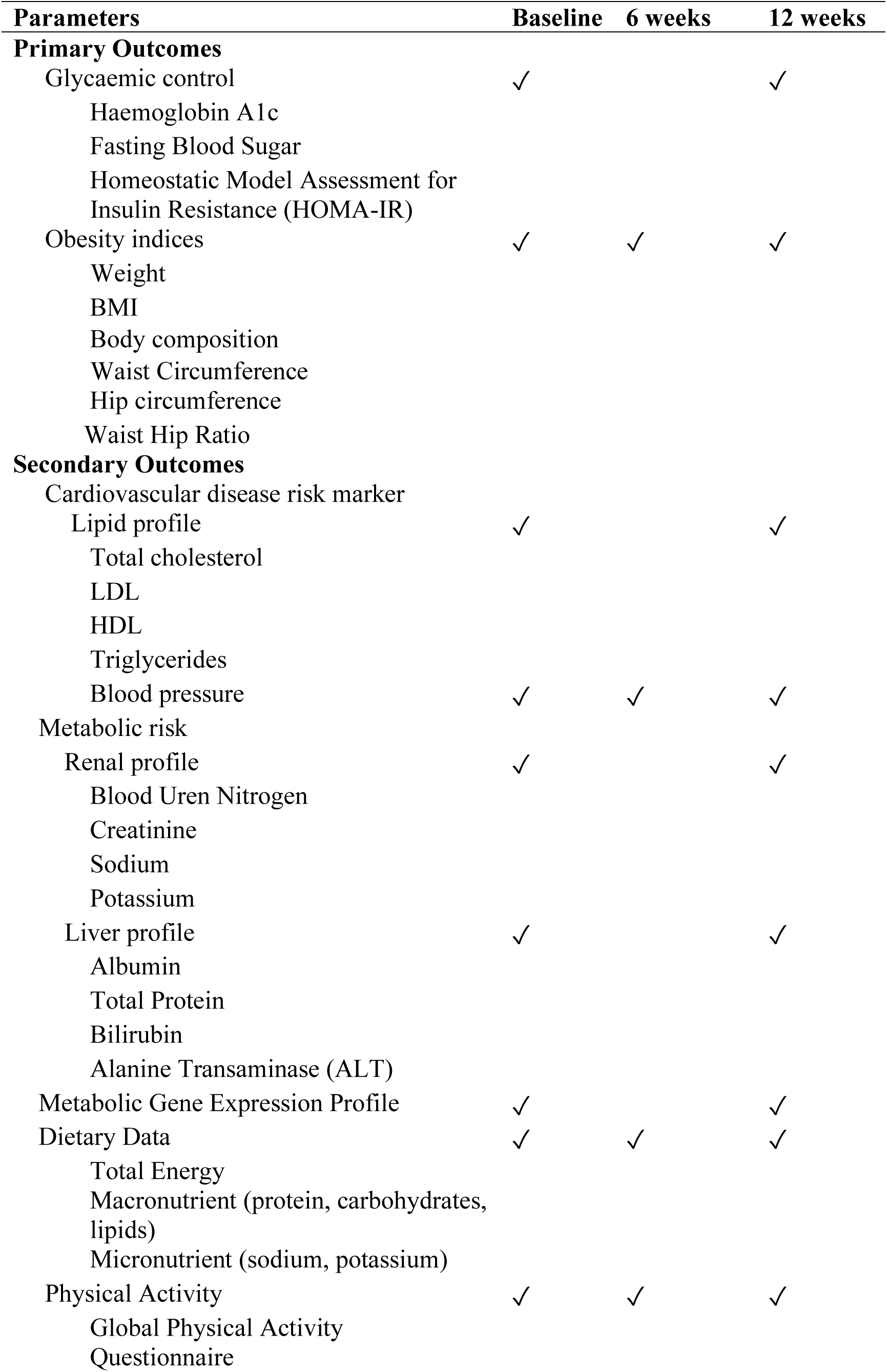
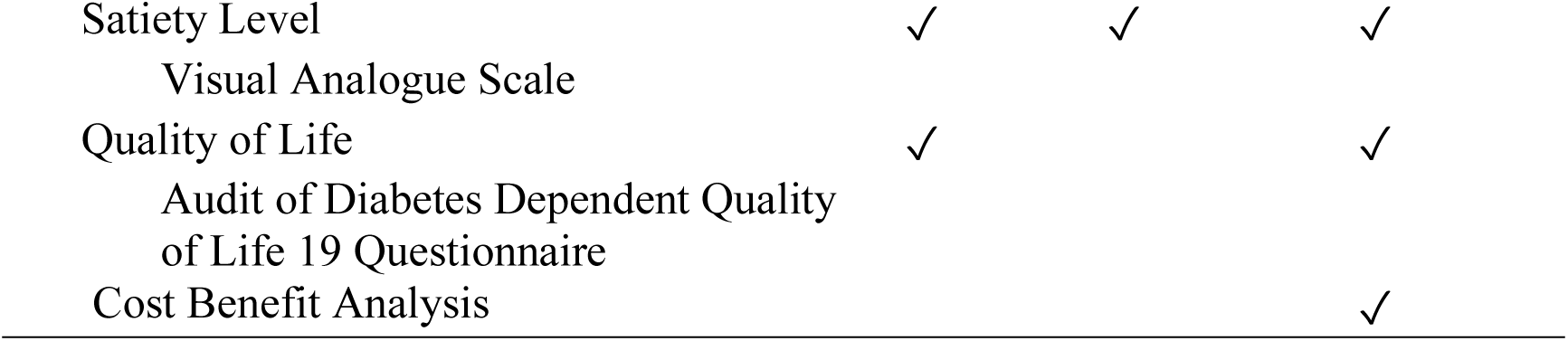
Summary of the Parameters.

### Intervention group

The intervention group will receive a meal replacement product along with one session of one-to-one dietary consultation. The dietary consultation will follow the standard Medical Nutrition Therapy Guidelines for T2DM, second edition [27]. The dietary consultation session will be held during the baseline data collection. The suggested calorie intake will be calculated based on their current BMI and will be emphasised during the dietary consultation session. Participants will be instructed to replace their two main meals with the MR product (Resurge DM, Quantum Upstream Sdn Bhd), consisting of 53 g of powder mixed with 200 ml of water. Instructions on how to prepare the MR will be provided individually. Nutritional information for the MR product is displayed in Table 2. Participants will also be given a weekly checklist form and an adverse effect form that comes together with the MR product. Information comes with instructions on how to complete the checklist. Participants allocated in the intervention group will be provided with 72 packets of MR products during their visit at the beginning and at week 6, together with detailed instruction on how to prepare them. During the first two weeks of the intervention, participants will also be supplied with oats and chia seeds to facilitate the transition from solid to liquid meals. Participants will mix a serving of oats and chia seed given together with the MR. A serving of oats will provide 75 kcal, while a serving of chia seeds will provide 34 kcal. This initial period will be considered a transition phase. The transition phase in this study will allow patients to adjust gradually to the new diet regimen. Besides, a gradual transition may reduce the likelihood of participants dropping out due to the difficulty in adapting to a new diet regimen. After two weeks of transition phase, participants will continue to replace two main meals with a serving of MR alone for the rest of the study. Study materials (checklist and adverse effect form) are returned on week 12. A visual analogue scale of satiety level will be distributed to the participants during the baseline visit since participants need to record it before and after taking the meal. A member of the study team will contact participants weekly to verify adherence to the dietary intervention and potential adverse events.

**Table 2.**
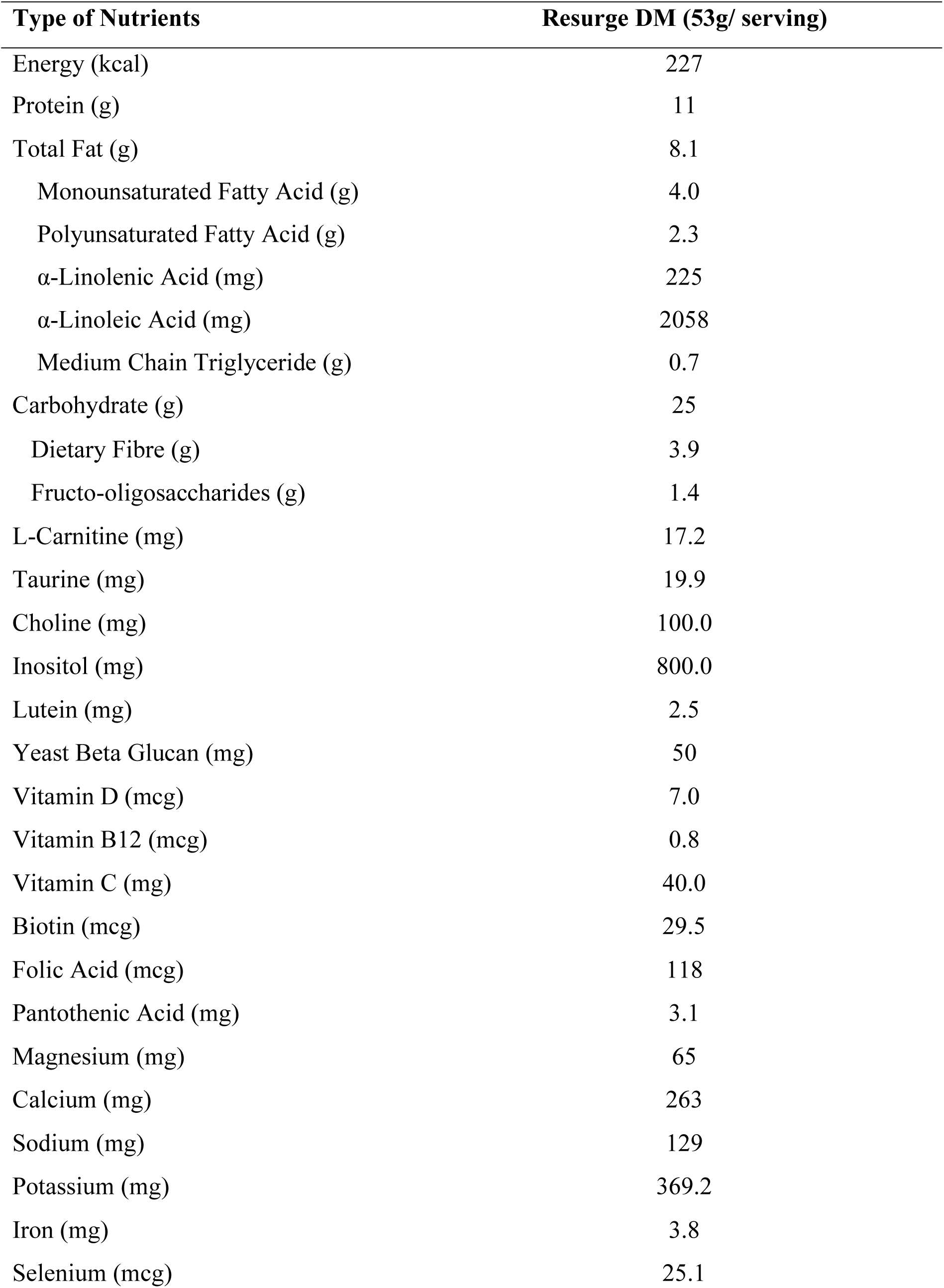
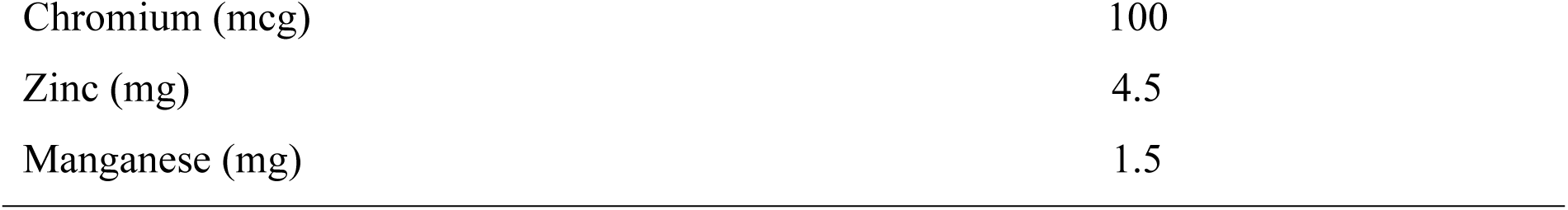
Nutritional information of a diabetes-specific meal replacement (MR)

### Control group

The control group will be provided with one, one to one dietary consultation session during the baseline collection. The dietary consultation will follow the standard Medical Nutrition Therapy for T2DM [27]. Participants’ suggested calorie will also be calculated based on their current BMI. A visual analogue scale of satiety level will also be distributed to the participants during the baseline visit since participants need to record it before and after their main meal intake.

## PRIMARY OUTCOME

### Glycaemic Control

Glycaemic control that will be measured in this study is HbA1c, fasting blood glucose, and fasting insulin levels. At baseline and 12th week follow-up sessions, participants will be asked to fast overnight for at least 10 hours for blood sample collection purposes. Peripheral vein blood samples will be drawn by a trained phlebotomist. A total of 14 ml of blood will be collected into specific tubes (plain tube, sodium fluoride tube, and ethylenediaminetetraacetic acid (EDTA) tube) and immediately stored in an ice box for delivery. The HbA1c and fasting blood glucose will be analysed at the medical laboratory of Hospital Canselor Tuanku Muhriz, National University of Malaysia. Meanwhile, the insulin resistance level will be sent to an outside lab (Lablink M Sdn. Bhd), since the test is not available in the medical laboratory of Hospital Canselor Tuanku Muhriz, National University of Malaysia. Based on the fasting blood glucose and fasting insulin level, the homeostatic model assessment for insulin resistance (HOMA-IR) will be calculated. The results of the blood test will be mailed or given to the participants during the next follow-up. Contact details are provided along with the results so that participants can contact the investigators or physicians if they have any questions.

### Obesity Indices

The obesity indices included weight, body mass index (BMI), body composition, waist circumference, hip circumference, and waist hip ratio (WHR). At the baseline, week 6 and week 12, anthropometric measurements are taken twice, and the average is used for data analysis. In light indoor clothes, the body weight will be measured in kilos using an electronic weighing scale, InBody 270 (kg). To avoid swaying or tipping, the scale will be set on a level, firm, and sturdy surface. Participants are requested to remove their socks and anything else from their bodies that may cause an inaccuracy in the measurements before they are collected. The weight will be calculated to the nearest 100 g (0.1 kg). Within 100 g, two measurements performed in quick succession should match (0.1 kg). Using the SECA 217 stadiometer, the participant’s height will be measured in centimetres. To eliminate parallax error, the measurements will be taken at eye level with the headboard and read to the nearest 0.1 cm. The BMI will be calculated based on the height and weight value according to a formula. For the categorisation, BMI will be categorised by using the Clinical Practice Guideline Management of Obesity [26] cut-off point as underweight (<18.5 kg/m2), normal (18.5-22.9 kg/m2), overweight (23-26.9 kg/m2), and obese (≥27.0 kg/m2). Next, for the waist circumference, it will be measured with a SECA measuring tape (SECA, Germany). The participants will be requested to stand up straight and breathe properly before beginning the measurement. Using fingers, the highest point of the hip bone and the bottom of the ribs will be found. The measuring tape is tight against the waist and put between the two spots. The spots are the highest point of the iliac crest and the last floating rib. At the conclusion of a regular expiry, the waist circumference is measured. Waist circumference individuals were at risk if their measurements were >90 cm for males and >80 cm for females [28]. For the hip circumference, the identical measuring tape is used (SECA measuring tape). The participants are asked to stand straight with arms at sides and feet together. With the measurer on his knee on the right side of the participants, the tape is held in a horizontal plane at the spot providing the highest circumference across the buttocks. WHR will be calculated based on the measurement taken and will be categorised based on gender, which is >0.90 for males and >0.85 for females [28]. Prior to the measurements, the equipment used in this study was calibrated. All anthropometric measurements are performed with subjects wearing light clothing without socks and shoes.

## SECONDARY OUTCOME

### Cardiovascular Risk Marker

In this study, the CVD risk markers are blood pressure and lipid profile. The ambulatory blood pressure will be measured using an electronic blood pressure machine with an appropriate cuff size (Omron Corporation, Kyoto, Japan). The participants’ blood pressure is assessed, and the mean readings are used. The blood pressure readings are taken from the seated participants’ left arm and will be taken twice. High blood pressure reading is defined when systolic blood pressure (SBP) >140 mmHg and/or diastolic blood pressure (DBP) >90mmHg [29]. Meanwhile, for the lipid profile, the blood that had been collected for the sugar profile will also be sent for lipid profile analysis, which consists of HDL, LDL, total cholesterol, and triglyceride level. The baseline and week 12 readings will be compared.

### Blood Profile

Other biochemical profiles that are looked at are triglycerides, HDL, LDL, total cholesterol, triglycerides, blood urea nitrogen, creatinine level, sodium, potassium, total protein, albumin, bilirubin, and alanine transaminase (ALT). The blood will be analyzed in the medical laboratory at Hospital Canselor Tunku Mukhriz, National University of Malaysia. The results of the blood test will be mailed or given to the participants during the next follow-up. Contact details are provided along with the results to enable the participants to contact the investigators or physician if they have any enquiries.

For the metabolic gene expression profile analysis, an additional 12 ml of fasting blood will be collected using an EDTA tube from 12 participants (6 from the control group and 6 from the intervention group) at baseline and 12th weeks. Peripheral blood mononuclear cells (PBMC) will be isolated from the blood sample following the protocol [30] and stored in cyrobox at -80°C prior to RNA extraction.

### Metabolic Gene Profile Analysis

Total RNA will be extracted from PBMC using the Blood/Cell RNA Mini Kit with DNase from Geneaid, following the protocol provided by the manufacturer. The extracted RNA samples will be treated with DNase to remove DNA contaminants. The RNA quality will then be determined using Nanodrop spectrophotometer. Pure RNA has a A260/A280 ratio of 1.9-2.1. All RNA samples will be stored at -80°C until gene expression analysis. All RNA samples will then be sent to the Genomax Technologies laboratory for gene expression analysis. This analysis is performed using the nCounter Human Metabolic Pathway by Nanostring following the protocol provided by the manufacturer. The nSolver software will be used to analyze the gene expression dataset to determine changes that occur to the metabolic gene networks and pathways.

### Dietary Data

At the beginning of the study, an experienced research dietitian will conduct dietary assessments for each participant. To monitor dietary intake throughout the study period, participants will be required to maintain a 3-day dietary record during baseline, week 6 and week 12 [31]. This record will document the food and beverages consumed by the participants over two weekdays and one weekend day within a week. Participants will be instructed to provide detailed information about their meals, including the types of foods consumed, quantities, preparation methods, brand names of food items, sauces, and ingredients used. To assist participants in estimating portion sizes accurately, they will be shown pictures of various household measurements based on the Atlas of Food Exchanges & Portion Sizes [32]. Additionally, any supplements, such as vitamins or minerals, consumed by the participants will also be recorded. The mean caloric and nutrient intake data collected from the dietary records will be analysed using the Nutritionist Pro software (Axxya Systems, Stafford, TX, USA) for computer-based analysis.

### Physical Activity

The physical activity levels of the participants are assessed by using a validated Malay version of the Global Physical Activity Questionnaire (GPAQ) for Malaysian population [33] will be employed. This questionnaire will be self-administered and if there are literacy issues, the questionnaire will be administered by the researcher. The GPAQ collects information about physical activity in three domains which are work, transport, and recreation, as well as average time per day spent in sedentary behaviour. The GPAQ is scored in minutes per day (min/d). Based on the guidelines provided by this questionnaire, the metabolic equivalent and total daily energy expenditure for each participant will be calculated using the MET value. This standardized instrument will enable the researchers to quantify the participants’ engagement in various physical activities and determine their overall energy expenditure levels.

### Satiety Level

In order to evaluate the perceived satiety of participants after consuming the meal replacement (in the treatment group) or a regular meal (in the control group), a Visual Analog Scale (VAS) questionnaire will be utilized [34]. Participants will be instructed to complete the VAS immediately before consuming their meal/meal replacement, and then again at specified intervals of 30 minutes, 60 minutes, 120 minutes, 180 minutes, and 240 minutes after consumption. An average appetite score will be calculated at each time point using a formula as describe and shown below [35]. This approach will allow the us to quantify and compare the subjective feelings of satiety experienced by participants across the two groups over an extended period following their respective meal or meal replacement consumption

Average appetite = desire to eat + hunger + (10−fullness) + prospective food consumption.

### Quality of Life

The study will use a validated Malay translation of the ADDQOL-19 questionnaire to evaluate the quality of life (QoL) of participants. This tool has demonstrated good validity and reliability in previous research [36]. The ADDQOL-19 comprises 21 items, out of which 19 items assess diabetes-dependent domains, while the remaining two items provide an overview of general aspects [37]. The questionnaire uses Likert scale which range from -3 to 3. The overall ADDQOL-19 score is calculated by combining the averages of the two domains and the equation is shown below. Higher score will indicate a better quality of life. Last but not least, to facilitate accurate responses, researchers will be available to assist participants in comprehending and completing the questionnaire items during its administration.

Overall ADDQOL-19 score = [(2 × Present life domain average) + Diabetes-dependent domain average] / 3

### Cost-Benefit

The cost-benefit analysis will be based on monetary value. The cost will be calculated based on the direct and indirect cost. The direct cost included the expenses of meal replacement and the labour cost of dietitian involved. Meanwhile, the benefit will be calculated based on the formula [38]. The formula consists of the number of the participants, the prevalence of T2DM, the percentage of the patient comply with the MR and the percentage of the participants that have reduce the hbA1c during the study. Then, the Net Present Value based on the formula shown below. The discount rate will be based on the average of the lending rate in Malaysia 2024 which is 5.35 % per annum [39].

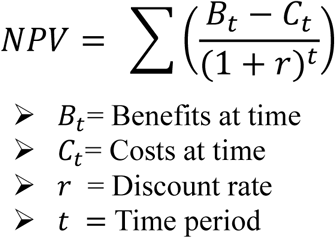

### Adverse Reaction Record

Based on the previous study [22], a few adverse events had been recorded, such as gastrointestinal discomfort, flatulence, and nausea. As a result, an adverse events form will be given to the participants at the start of the study, and those who experienced adverse events must report them immediately. Those who experienced an adverse event can receive treatment at Klinik Primer.

## ADHERENCE AND WITHDRAW/DISCONTINUATION

Participants are promptly removed from the research if they: (1) have a serious adverse effect, (2) get pregnant, (3) initiate or modify the use of drugs or supplements mentioned in the eligibility criteria, or (4) no longer fulfil the inclusion criteria. Participants in the MR group are required to bring back all MR bags, whether empty or not, during the visits on week 6 and 12. The bags used in the trial are calculated, and participants are eliminated from the study if there is more than 30% of the product remaining inside the bags. Furthermore, participants have the option to discontinue their involvement in the study at any given moment.

## STATISTICAL ANALYSES

### Sample size estimates

The sample size is calculated by using G *Power 3.1.4 software using mean difference value of the HbA1c; effect size 0.2055067 [40]. The protocol for calculation is as shown below in Table 3. Based on the calculation, a total of 126 participants will be needed for the study. In order to account for up to a 30% dropout, a total of 164 participants (82 in each group) will be recruited for this study.

**Table 3.**
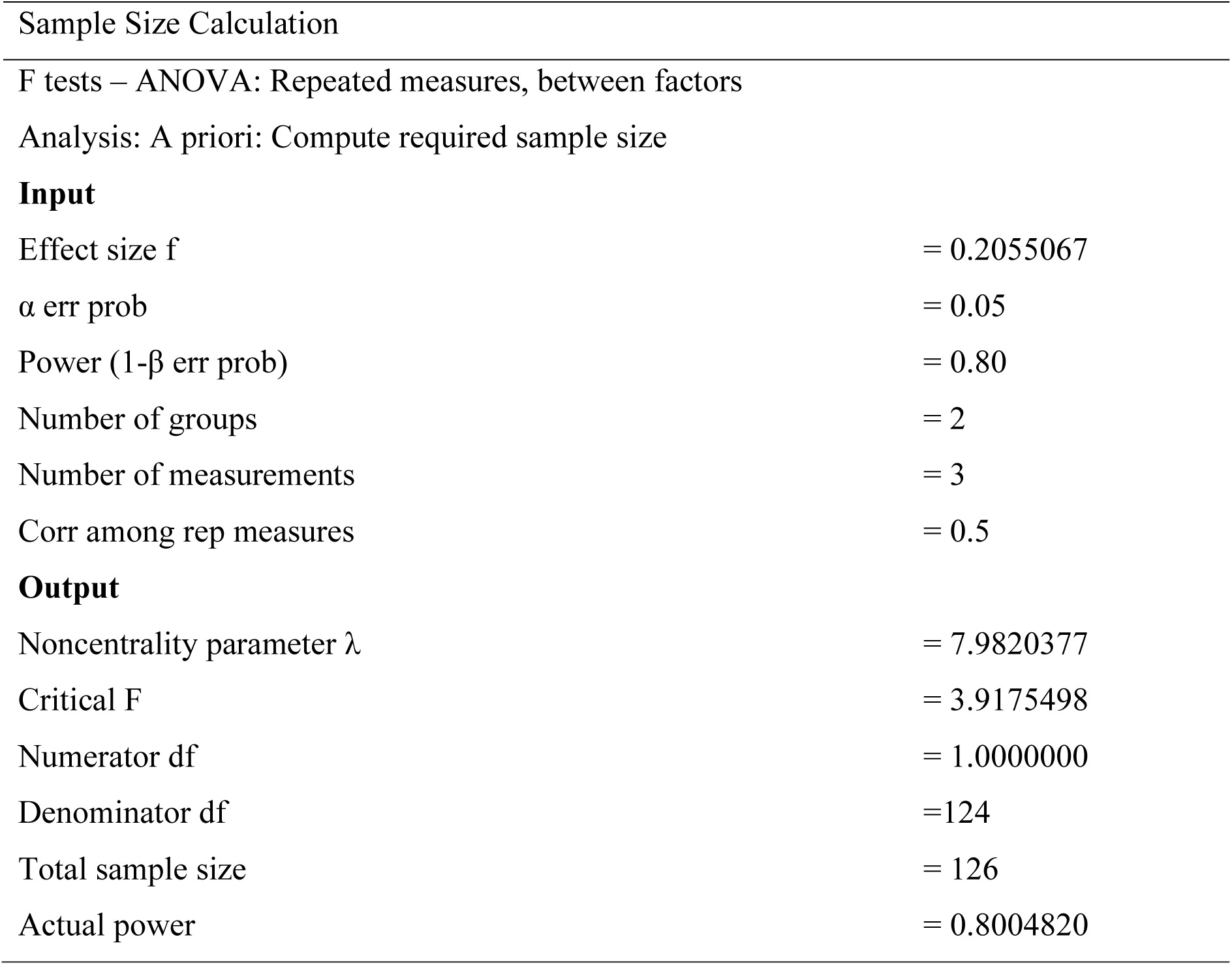
Protocol for calculation in G* Power.

### Data analysis

This is an intervention study that was aimed at determining the efficacy of partial diabetes-specific meal replacement on weight reduction, glycaemic control and metabolic gene expression among overweight and obese T2DM patients. All of the statistical analyses will be performed by using the IBM Statistical Package for the Social Sciences (SPSS) version 29.0 software with a significance level of p<0.05. Normality of data will be analysed by using the Shapiro-Wilk test and significant value p>0.05 which indicates normal distribution. The mean differences of the baseline data between the intervention and control group will be analysed using the Independent-t or Mann Whiteney test for continuous parameters.

On the other hand, Chi-Square was selected for the analysis of categorical variables. The effects of Resurge DM over time, the effect of group and its interaction will be determined by using mixed design repeated measures ANOVA. Mauchly’s test of Sphericity will be used to analyse the assumption sphericity for each dependent variable. For the variables with p≤0.05, will be indicated as a violation of this assumption and Greenhouse-Geisser adjustment will be made to the appropriate degree of freedom. Indication of differences within and between the periods of supplement consumption will be analysed using Bonferroni’s post hoc test. Sub-group analysis will be conducted if required.

## DISCUSSION

This randomised controlled trial is to assess the efficacy of a diabetes-specific meal replacement (MR) product in the management of type 2 diabetes mellitus (T2DM) in overweight and obese individuals T2DM patients. This study will take a comprehensive approach to managing T2DM by considering a few outcomes such as weight loss, control blood sugar levels, blood pressure, satiety level, quality of life, and changes in the gene expression profile. The incorporation of cost-benefit analysis additionally will help to improves the practical applicability of the results that will provide a valuable perspective for healthcare practitioners and policymakers especially in Malaysia [41]. In addition, by identify the metabolic gene expression profile it will clarify the molecular pathways that are responsible for the effects of the intervention, thereby enhancing the scientific comprehension of T2DM [42,43].

Nevertheless, the study has certain constraints that require attention. The accuracy of compliance measurement, which relies on the returns of MR containers, may be compromised, hence potentially undermining the validity of the adherence data. Moreover, the dependence on self-reported assessments for nutritional consumption, physical activity, and satiety level will brings about the potential for recall bias and mistakes. The sample size, while sufficient for an initial inquiry, may restrict the ability to apply the findings to a broader population. Although there are several limitations, the study’s results are anticipated to offer crucial insights into the efficacy of meal replacement treatment in managing type 2 diabetes.

The potential enhancements in glycaemic management, weight reduction, and quality of life have the ability to greatly improve clinical practice and patient outcomes. The study’s identification of alterations in metabolic gene expression may facilitate the development of personalised dietary therapies according to individuals’ genetic profiles. Ultimately, this study has the capacity to provide a valuable foundation of evidence supporting the utilisation of diabetes-specific meal replacement products in the management of Type 2 Diabetes Mellitus (T2DM). It has the potential to provide valuable insights for the creation of focused interventions that enhance the well-being of individuals with T2DM based on the factors that associated with T2DM and decrease the overall impact of this condition on public health worldwide [44]. It is advisable to conduct additional research with extended observation periods and larger groups of participants in order to confirm and build upon these findings [21].

## ETHICS AND DISSEMINATION

This study is approved by the National University of Malaysia’s Human Research Ethics Committee (JEP-2024-695). This study is registered on International Standard Randomised Controlled Trial Number (ISRCTN57040303) and in accordance with Good Clinical Practice Guidelines and the ethical principles of the Declaration of Helsinki 1964. All personal information will be kept private, and participation is anonymous. Participants are assigned a study ID, which is kept separated from any personal information collected. A master list with identifiable information and study IDs is protected and stored at the Centre for Health Ageing and Wellness (H-CARE). If participants withdraw consent, they are asked for permission to use the data collected until that point; however, if they deny it, their data is destroyed. Following data collection, analysis and review of findings, manuscripts will be prepared for submission to peer--reviewed journals and results presented in national and international conferences. Study findings will also be disseminated through social media. Data will be published regardless of outcomes and the National University of Malaysia retains the right to publish.

## Data Availability

All data produced in the present study are available upon reasonable request to the authors

## Contributors

All authors made substantial contributions to the conception or design of the protocol, data acquisition, and analysis or interpretation of data. All authors took part in drafting and revising the protocol critically, gave final approval for this final version to be published, and agreed to be held accountable for all aspects of the work. WANCWM, SS, LMT, RR and MI were involved in the design of the study. WANCWM, SS and MI wrote the study protocol. WANCWM, SS, LMT, RR, and MI participated in drafting and revising the manuscript. WANCWM, SS, LMT, RR, and MI read and approved the final manuscript.

## Funding

This work was supported by an industrial grant from Quantum Upstream Sdn Bhd (Kuala Lumpur, Malaysia). Grant number: NN-2023-019. Per contractual agreement, the funder has had no role in the study design and implementation, writing of the manuscript and decision to submit the article for publication.

## Competing interests

All authors declare no conflict of interest.

## Patient and public involvement

Patients and/or the public were not involved in the design, or conduct, or reporting, or dissemination plans of this research.

## Patient consent for publication

Not applicable.

## Ethics approval

JEP-2024-695

## Provenance and peer review

Not commissioned; externally peer reviewed.

## Supplemental material

This content has been supplied by the author(s). It has not been vetted by BMJ Publishing Group Limited (BMJ) and may not have been peer--reviewed. Any opinions or recommendations discussed are solely those of the author(s) and are not endorsed by BMJ. BMJ disclaims all liability and responsibility arising from any reliance placed on the content. Where the content includes any translated material, BMJ does not warrant the accuracy and reliability of the translations (including but not limited to local regulations, clinical guidelines, terminology, drug names and drug dosages), and is not responsible for any error and/or omissions arising from translation and adaptation or otherwise.

## Notes

### Competing Interest Statement

The authors have declared no competing interest.

### Funding Statement

This study was funded by Quantum Upstream Sdn.Bhd.

### Author Declarations

Ethical Committee National University of Malaysia (JEP-2024-695)

