## Supplementary material for "Evaluating Diabetes-Specific Meal Replacements for Glycaemic Control in Overweight and Obese T2DM Patients: A Protocol for a Randomized Controlled Trial": Table 1

Table 1 Summary of the Parameters

| Parameters | Baseline | 6 weeks | 12 weeks |
|---|---|---|---|
| **Primary Outcomes** | | | |
| Glycaemic control | ✓ | | ✓ |
| Haemoglobin A1c | | | |
| Fasting Blood Sugar | | | |
| Homeostatic Model Assessment for Insulin Resistance (HOMA-IR) | | | |
| Obesity indices | ✓ | ✓ | ✓ |
| Weight | | | |
| BMI | | | |
| Body composition | | | |
| Waist Circumference | | | |
| Hip circumference | | | |
| Waist Hip Ratio | | | |
| **Secondary Outcomes** | | | |
| Cardiovascular disease risk marker | | | |
| Lipid profile | ✓ | | ✓ |
| Total cholesterol | | | |
| LDL | | | |
| HDL | | | |
| Triglycerides | | | |
| Blood pressure | ✓ | ✓ | ✓ |
| Metabolic risk | | | |
| Renal profile | ✓ | | ✓ |
| Blood Uren Nitrogen | | | |
| Creatinine | | | |
| Sodium | | | |
| Potassium | | | |
| Liver profile | ✓ | | ✓ |
| Albumin | | | |
| Total Protein | | | |
| Bilirubin | | | |
| Alanine Transaminase (ALT) | | | |
| Metabolic Gene Expression Profile | ✓ | | ✓ |
| Dietary Data | ✓ | ✓ | ✓ |
| Total Energy | | | |
| Macronutrient (protein, carbohydrates, lipids) | | | |
| Micronutrient (sodium, potassium) | | | |
| Physical Activity | ✓ | ✓ | ✓ |
| Global Physical Activity Questionnaire | | | |
| Satiety Level | ✓ | ✓ | ✓ |
| Visual Analogue Scale | | | |

| | | |
|---|---|---|
| Quality of Life | ✓ | ✓ |
| Audit of Diabetes Dependent Quality of Life 19 Questionnaire | | |
| Cost Benefit Analysis | | ✓ |

Table 2 Nutritional information of a diabetes-specific meal replacement (MR)

| Type of Nutrients | Resurge DM (53g/ serving) |
|---|---|
| Energy (kcal) | 227 |
| Protein (g) | 11 |
| Total Fat (g) | 8.1 |
| Monounsaturated Fatty Acid (g) | 4.0 |
| Polyunsaturated Fatty Acid (g) | 2.3 |
| α-Linolenic Acid (mg) | 225 |
| α-Linoleic Acid (mg) | 2058 |
| Medium Chain Triglyceride (g) | 0.7 |
| Carbohydrate (g) | 25 |
| Dietary Fibre (g) | 3.9 |
| Fructo-oligosaccharides (g) | 1.4 |
| L-Carnitine (mg) | 17.2 |
| Taurine (mg) | 19.9 |
| Choline (mg) | 100.0 |
| Inositol (mg) | 800.0 |
| Lutein (mg) | 2.5 |
| Yeast Beta Glucan (mg) | 50 |
| Vitamin D (mcg) | 7.0 |
| Vitamin B12 (mcg) | 0.8 |
| Vitamin C (mg) | 40.0 |
| Biotin (mcg) | 29.5 |
| Folic Acid (mcg) | 118 |
| Pantothenic Acid (mg) | 3.1 |
| Magnesium (mg) | 65 |
| Calcium (mg) | 263 |
| Sodium (mg) | 129 |
| Potassium (mg) | 369.2 |
| Iron (mg) | 3.8 |
| Selenium (mcg) | 25.1 |
| Chromium (mcg) | 100 |
| Zinc (mg) | 4.5 |

| | |
|---|---|
| Manganese (mg) | 1.5 |

Table 3 Protocol for calculation in G* Power.

| Sample Size Calculation | |
|---|---|
| F tests – ANOVA: Repeated measures, between factors | |
| Analysis: A priori: Compute required sample size | |
| **Input** | |
| Effect size f | = 0.2055067 |
| α err prob | = 0.05 |
| Power (1-β err prob) | = 0.80 |
| Number of groups | = 2 |
| Number of measurements | = 3 |
| Corr among rep measures | = 0.5 |
| **Output** | |
| Noncentrality parameter λ | = 7.9820377 |
| Critical F | = 3.9175498 |
| Numerator df | = 1.0000000 |
| Denominator df | =124 |
| Total sample size | = 126 |
| Actual power | = 0.8004820 |
